# Geographic clusters of objectively measured physical activity and the characteristics of their built environment in a Swiss urban area

**DOI:** 10.1101/2021.04.23.21255966

**Authors:** Juan R Vallarta-Robledo, Stéphane Joost, Marco André Vieira Ruas, Cédric Gubelmann, Peter Vollenweider, Pedro Marques-Vidal, Idris Guessous

## Abstract

**Introduction:** Evidence suggests that the built environment can influence the intensity of physical activity. However, despite the importance of the geographic context, most of the studies do not consider the spatial framework of this association. We aimed to assess individual spatial dependence of objectively measured moderate and vigorous physical activity (MVPA) and describe the characteristics of the built environment among spatial clusters of MVPA.

**Methods:** Cross-sectional data from the second follow-up (2014-2017) of CoLaus|PsyCoLaus, a longitudinal population-based study of the Lausanne area (Switzerland), was used to objectively measure MVPA using accelerometers. Local Moran’s I was used to assess the spatial dependence of MVPA and detect geographic clusters of low and high MVPA. Additionally, the characteristics of the built environment observed in the clusters based on raw MVPA and MVPA adjusted for socioeconomic and demographic factors were described.

**Results:** Data from 1,889 participants (median age 63, 55% women) were used. The geographic distribution of MVPA and the characteristics of the built environment among clusters were similar for raw and adjusted MVPA. In the adjusted model, we found a low concentration of individuals within spatial clusters of high MVPA (median: 36.9 mins; 3% of the studied population) and low MVPA (median: 10.1 mins; 2% of the studied population). Yet, clear differences were found in both models between clusters regarding the built environment; high MVPA clusters were located in areas where specific compositions of the built environment favor physical activity.

**Conclusions:** Our results suggest the built environment may influence spatial patterns of MVPA independently of socioeconomic and demographic factors. Interventions in the built environment should be considered to promote physically active behaviors in urban areas.

## Introduction

Regular physical activity improves health status and prevents chronic diseases [1,2]. For such reasons, the World Health Organization (WHO) recommends 150 minutes of moderate-intensity, 75 minutes of vigorous-intensity per week, or a combination of moderate and vigorous physical activity (MVPA) [3]. However, despite the benefits of consistent MVPA, current trends do not show an encouraging increment of these recommendations in the population [4]. Regular practice of physical activity depends on a series of determinants, including biological, psychological, cultural, socioeconomic, and environmental [5,6]. The built environment can influence physical activity and promote healthy lifestyles through different settings [7,8]. Indeed, individuals living in areas with higher accessibility to walking, cycling, and public transport infrastructure, street connectivity and accessibility to diverse destinations, the presence of diverse land use (i.e. residential zones mixed with public, recreational and commercial areas), larger access to recreational areas of quality, and greater population density have been associated with higher levels of physical activity [9–11].

This is of major importance as it supports the development of public policies promoting physically active behaviors at the community level. To facilitate healthy environments and target areas for intervention, it is important to identify and understand the spatial distribution of health outcomes and environmental characteristics [12,13]. Spatial statistics and related clustering methods are of great help to reveal non-random geographic patterns of a given outcome and have become popular in recent years in epidemiological studies [14,15]. However, despite the importance of the geographic context in the association between the built environment and physical activity, most previous research did not consider the spatial factor.

Studies using a spatial methodology to assess this relationship have shown partial associations [16–23]. This mixed evidence may result from the fact that these works analyze the association at an aggregated level (i.e. statistical subsectors, counties, etc.) [16– 20], which may not always reflect the real geographic context [24,25]. Furthermore, of these studies, only one objectively measured physical activity [18]. Among the research works assessing spatial patterns of physical activity at an individual scale, all were based on self-reported physical activity [21–23], which limits their conclusions.

We aimed to assess the spatial dependence of objectively measured MVPA in the urban area of Lausanne using individual accelerometry and geolocated data. Additionally, we measured and described the built environment characteristics among the spatial clusters of both raw MVPA and MVPA adjusted for socioeconomic and demographic factors.

## Materials and Methods

### Urban area of Lausanne

The city of Lausanne covers an area of 41.37 km^2^ with a population of 144,790 inhabitants in 2017, and it is divided into 81 statistical sub-sectors (https://www.lausanne.ch/officiel/statistique.html). It has around 350 hectares of parks and gardens, 111 km of cycling pathways (https://www.lausanne.ch/officiel/statistique.html), two metro and 42 bus and trolleybus lines (https://rapportannuel.t-l.ch) (S1 Fig).

### Health data

Data were obtained from CoLaus|PsyCoLaus cohort, a longitudinal population-based study started in 2003-2006, whose main aim is to explore the determinants of cardiovascular diseases in individuals aged 35-75 years in Lausanne, Switzerland [26]. The second follow-up of the cohort was carried out from 2014 to 2017 and collected physical activity information by accelerometry (see below). Because of this, we only considered cross-sectional data from this second follow-up for analysis.

Only individuals of the second follow-up who participated in the collection of accelerometry data were considered for analysis. Participants were excluded if they did not have valid accelerometry data (see below), lived of outside the urban area of Lausanne, or if geolocated data or covariates (see below) were missing.

The institutional Ethics Committee of the University of Lausanne, which afterwards became the Ethics Commission of Canton Vaud (www.cer-vd.ch) approved the baseline CoLaus study (reference 16/03). The approval was renewed for the first (reference 33/09) and the second (reference 26/14) follow-ups. The study was performed in agreement with the Helsinki declaration and its former amendments, and in accordance with the applicable Swiss legislation. All participants gave their signed informed consent before entering the study.

### Physical activity data collection process

A detailed description of the physical activity data collection process was described previously by Gubelmann et al. [27]. Overall, physical activity data were obtained from a wrist-worn triaxial accelerometer (GENEActiv, Activinsights Ltd., United Kingdom) used in the right wrist of the participants at a frequency of 50 Hz for 14 days and converted into 1-minute epoch files using the GENEActiv macros [28]. The GGIR algorithm [29] was then used to transform accelerometry data suitable for analysis. Additionally, we used the PAMPRO methodology [30] as a sensitivity analysis to validate the findings obtained, results are shown in S2 Fig. Data were valid if individuals had accelerometry information ≥10 h during weekdays and ≥8 h during weekends [27]. A minimum of 5 days for weekdays and 2 days for weekends of accelerometry recorded data were required to consider observations valid for analysis [31].

Daily sedentary, light, moderate, and vigorous physical activity intensities were averaged and stratified by weekdays and weekends. MVPA intensities were considered to assess physical activity. MVPA is defined as the sum (in minutes) of the average daily moderate and vigorous physical activity obtained from the accelerometry data. Stratified results for weekdays and weekends are shown in S3 Fig.

### Covariates

Individual data were self-reported and physical measures (i.e. height and weight) were taken by trained professionals in a single visit at the Centre Hospitalier Universitaire Vaudois. Socioeconomic and demographic covariates were age (years), gender, ethnicity (white and non-white), marital status (single, married, divorced, or widowed), education level (low, medium, or high), and work status (low, medium, high, or not working). We also included the annual median household income at the neighborhood level (1 CHF= 1.10 USD, November 2020) obtained from the 2009 Lausanne Census (https://www.vd.ch/themes/etat-droit-finances/statistique) and assigned to the place of residence of each individual. Additionally, we assessed the body mass index (BMI). As factors characterizing the built environment, we considered the number of parks and their location obtained from the Federal Office of Topography (https://www.swisstopo.admin.ch/en/home.html), cycling pathways length and preferential pedestrian zones (pedestrian and meeting places) from the Service des Routes et de la Mobilité (https://www.asitvd.ch/), population density (years 2014-2017) and land use mix area coverage from the Federal Statistical Office (FSO) reported at the hectometric level (https://www.bfs.admin.ch/bfs/en/home.html). Land use mix area coverage was calculated using the formula proposed by Frank et al. [32] and adapted to 5 land use categories reported by the FSO (residential, commercial & industrial, public, recreational, and natural). We also used this formula to calculate the coverage area of each individual land use category. Results range from 0 to 1; 0 stands for no land use mix area coverage and 1 for maximum land use mix area coverage. Likewise, to evaluate public transport accessibility, we calculated the number of public transport stops and the walking time it takes to reach the closest public transport stop from the place of residence of each individual using street network data from © OpenStreetMap contributors (https://www.openstreetmap.org/copyright) and Tobler’s Hiking function [33]. We also used this data source to calculate street connectivity (3 or greater number of streets intersections). All these built environmental variables were assigned to the place of residence of each participant within a buffer of 800 m and, except for land use mix area coverages and walking time to reach the closest public transport stop, values were divided by the buffer’s area (1.98 km^2^).

### Statistical analysis

We calculated Local Moran’s I (LMI) to identify possible spatial clusters of MVPA. LMI measures spatial dependence and highlights local clusters of the variable of interest, here MVPA [34]. This method was derived from the global Moran’s I index that evaluates spatial autocorrelation of a variable across a geographic area and ranges from −1 to 1 [35]. A value of 0 indicates no spatial dependence (observations are randomly distributed in space). A Moran’s scatterplot displays the relationship between the observed MVPA for individual i and the mean MVPA for the individuals located in a specified neighborhood around individual i (the weighted MVPA). Four distinct categories are identified according to the position of individuals in the four quadrants of the Moran’s scatterplot: i) high-high: individuals with a high MVPA surrounded by neighbors showing a high MVPA also, ii) low-low: individuals with a low MVPA surrounded by neighbors with a low MVPA also, iii) low-high: individuals with low MVPA surrounded by neighbors with a high MVPA (considered as outliers), iv) high-low: individuals with a high MVPA and surrounded by neighbors with low a MVPA (outliers also). There is an additional fifth category corresponding to individuals which are randomly distributed in space. The latter category is determined using a significance test based on Monte-Carlo random permutations of MVPA observations [34]. It is calculated as (M+1) / (P+1); P corresponds to the number of permutations and M to the number of instances where a permutation statistic is greater or equal than the observed value (if positive LMI), or lower or equal than the observed value (if negative LMI). In this study, we tested 999 Monte-Carlo permutations and an α level of 0.05, the spatial lag used was 800 m [36]. LMI was calculated on raw MVPA and on MVPA adjusted for socioeconomic and demographic factors using a linear median regression (S1 Appendix). MVPA exhibited a non-normal distribution and was transformed to its square root to obtain a normal distribution for statistical analysis and was back-transformed to its real value for the description of the results. We also ran LMI using different spatial lags of 400, 600, 1000, and 1200 m, and the analyses showed similar spatial patterns to the ones we obtained with 800m (S4 Fig).

Due to the low number of individuals contained within the spatial clusters observed, statistical comparisons were carried out using non-parametric methods (significance threshold of p<0.05); chi-square or fisher tests (when N < 5) for categorical data, and Kruskal Wallis tests (including Bonferroni-Holm’s correction) for numeric variables. Because of the above, data are reported as median and interquartile range (IQR) for numeric variables and frequencies and percentages (%) for categorical information. Analyses were performed in R version 3.6.3 [37]. Additionally, the rgeoda library [38] was used to run spatial analyses, sf [39] to calculate densities and lengths in spatial buffers for each individual. Maps were drawn with ggplot2 [40].

## Results

The initial sample consisted of 2,967 individuals that participated in the collection of accelerometry data. Among them, 945 individuals living outside the urban area of Lausanne and without valid accelerometry and geolocation data were removed, leading to a dataset of 2,022 observations (68%). A further 133 (6%) observations had missing data and were also removed, leading to a final sample of 1,889 individuals.

Individual variables and characteristics on the built environment of the studied and removed individuals are described in S1 Table. Overall, the median age of the studied individuals was 63 years (IQR: 16) and there were 1,409 (55%) women. The mean daily MVPA was 20.3 mins (IQR: 23.0).

### Spatial clusters of raw MVPA and adjusted MVPA

We detected spatial clusters of raw MVPA in the urban area of Lausanne (Fig 1). We identified one small, but statistically significant, concentration of individuals (1%) in clusters of high MVPA and a concentration of 2% of participants with discordant MVPA levels (Low-High) in the south-western region of the urban area (landmark #1). A small concentration (1%) of participants in low MVPA clusters and individuals having intertwined values of MVPA (High-Low) were observed in the north-western, eastern, and southern regions (landmarks #2-4).

**Fig 1.**
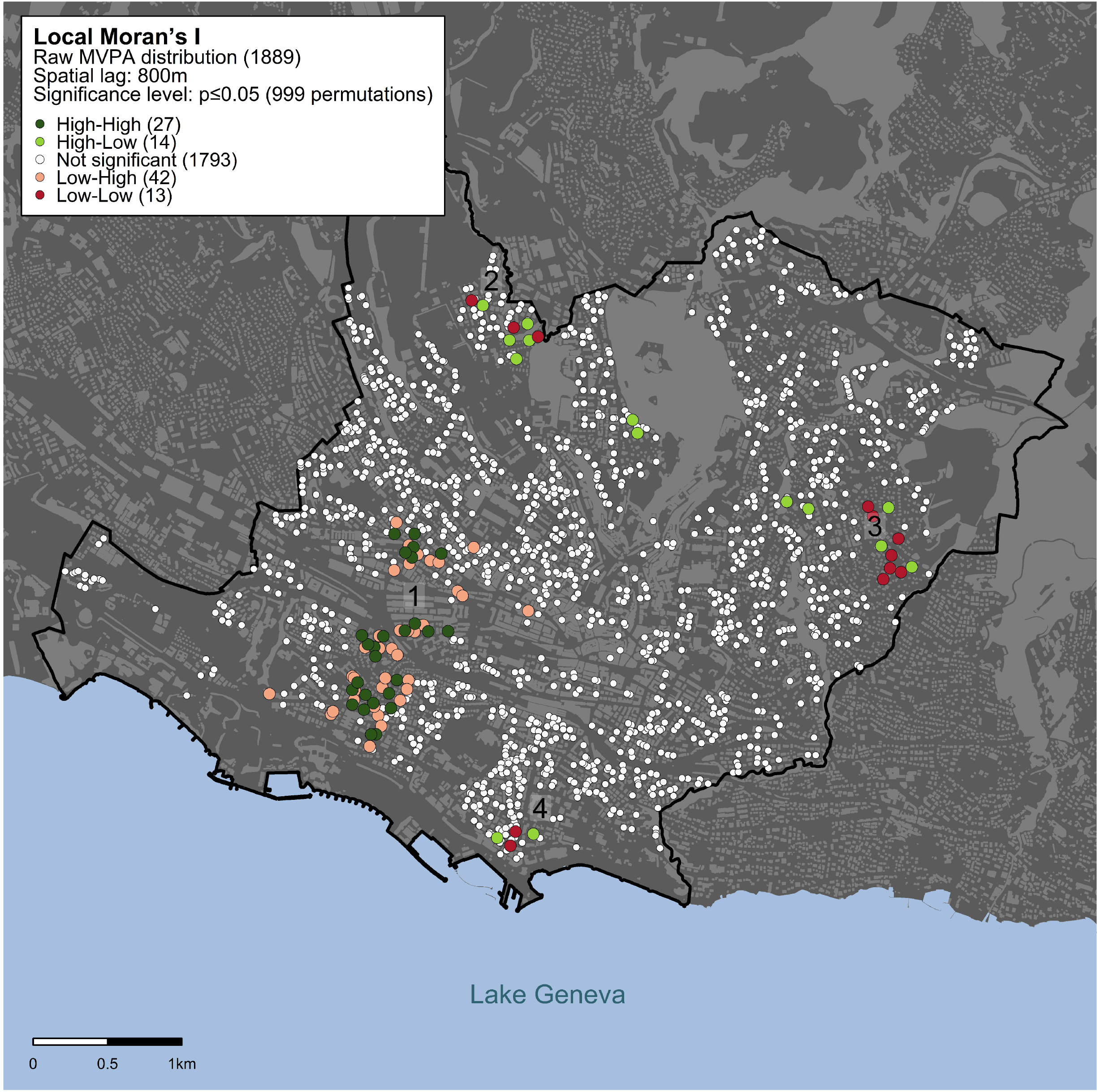
Spatial clusters of raw MVPA using Local Moran’s I statistics. Statistical significance is assessed based on an α threshold of p<0.05 within a spatial lag of 800 m. Dark-green dots indicate individuals with high MVPA values surrounded by neighbors also showing high MVPA values; red dots indicate individuals with low MVPA surrounded by neighbors with low MVPA values; light-green dots indicate individuals with high MVPA values surrounded by neighbors showing low MVPA values; pink dots indicate individuals with low MVPA values surrounded by neighbors with high MVPA values; white dots indicate individuals whose MVPA values are randomly distributed in the geographic space. Landmarks (1-4) are shown to facilitate the description and interpretation of the results.

Spatial clusters of MVPA adjusted for socioeconomic and demographic factors (Fig 2) were geographically distributed similarly to the clusters of raw MVPA (Fig 1). However, there was a higher concentration of Low-Low and High-Low clusters in the north-western area (landmark #2) and a lower concentration in the eastern and southern regions (landmarks #3 and #4). The overall size of low-low clusters increased from 1% to 2% of the studied population. High-High clusters increased their size from 1% to 3% (landmark #1).

**Fig 2.**
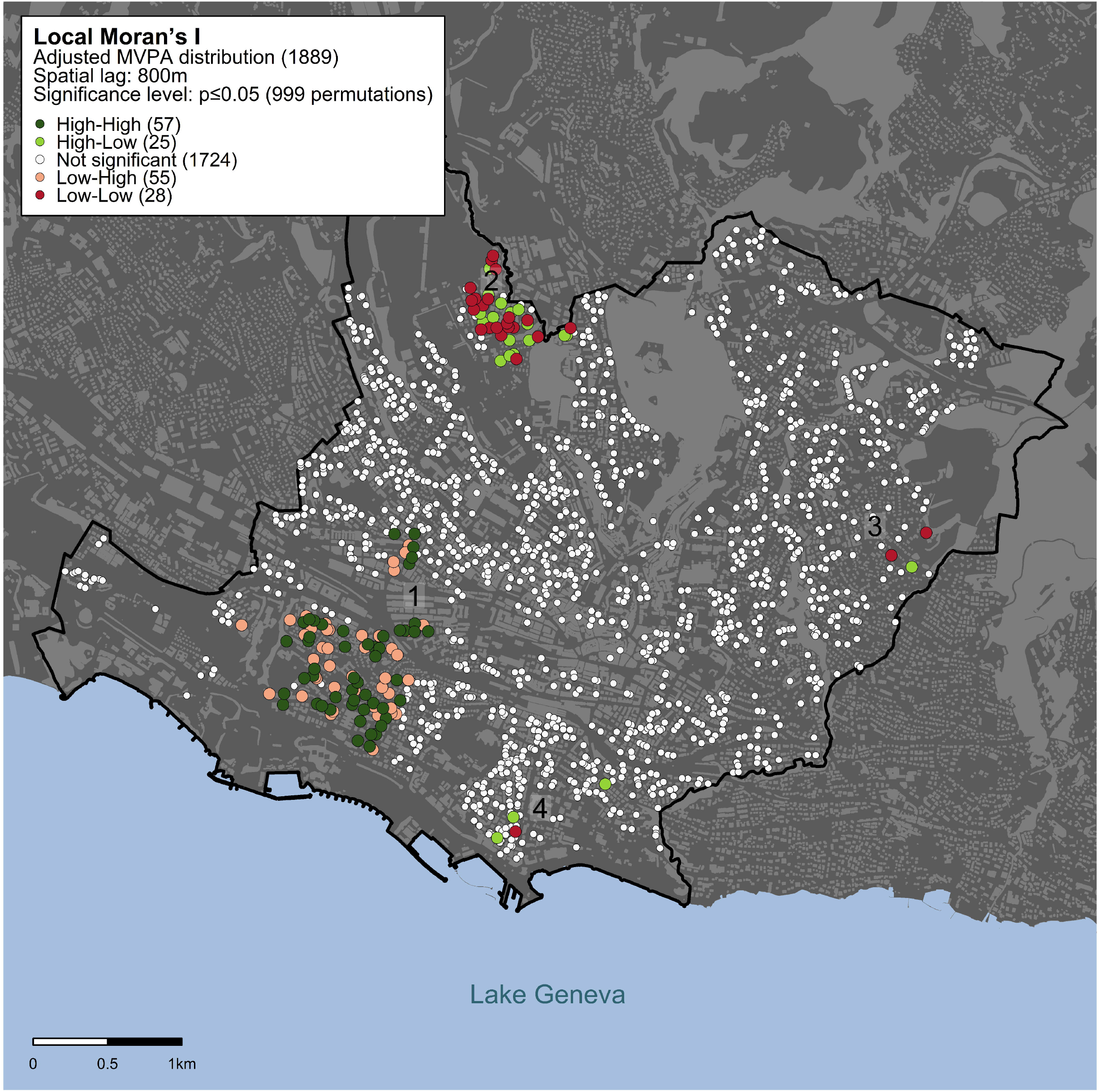
Spatial clusters of MVPA adjusted for socioeconomic and demographic factors using Local Moran’s I statistics. Statistical significance is assessed based on an α threshold of p<0.05 within a spatial lag of 800 m. Dark-green dots indicate individuals with high MVPA values surrounded by neighbors also showing high MVPA values; red dots indicate individuals with low MVPA surrounded by neighbors with low MVPA values; light-green dots indicate individuals with high MVPA values surrounded by neighbors showing low MVPA values; pink dots indicate individuals with low MVPA values surrounded by neighbors with high MVPA values; white dots indicate individuals whose MVPA values are randomly distributed in the geographic space. Landmarks (1-4) are shown to facilitate the description and interpretation of the result.

### Built environment characteristics in raw MVPA spatial clusters

Statistically significant built environment differences (p<0.05) were observed among spatial clusters of raw MVPA (Table 1). Clusters with high levels of MVPA (High-High) were located in areas with a higher number of parks (high: 2 *vs* low: 0), public transport stops (high: 28 *vs* low: 11), preferential pedestrian areas (high: 3 *vs* low: 2), cycling pathways (high: 6.7 *vs* low: 1.8 km), interconnected streets (high: 133 *vs* low: 55), population density (high: 11,708 *vs* low: 5,061), and public (high: 0.28 *vs* low: 0.21), and commercial & industrial land use areas (high: 0.18 *vs* low: 0.03) compared to clusters of low MVPA (Low-Low). In contrast, clusters of low MVPA were in locations with a higher presence of natural areas (low: 0.25 *vs* high: 0). Socioeconomic characteristics of individuals among clusters can be observed in S2 Table; older participants and the proportion of non-workers were statistically higher in low MVPA clusters.

**Table 1.** Built environment characteristics in raw MVPA spatial clusters.

| Built environment characteristics | No spatial dependence | High-High | Low-Low | High-Low | Low-High | p-value<sup>a</sup> |
| --- | --- | --- | --- | --- | --- | --- |
| N | 1793 (95%) | 27 (1%) | 13 (1%) | 14 (1%) | 42 (2%) |  |
| MVPA (mins) | 20.3 (23.2) | 35.2 (29.9) | 16.5 (9.8) | 37.8 (26.1) | 12.1 (10.0) | <0.001<sup>b</sup> |
| Number of parks | 2 (2) | 2 (1) | 0 (1) | 0 (1) | 2 (1) | <0.001<sup>c</sup> |
| Number of public transport stops | 20 (10) | 28 (5) | 11 (4) | 16 (4) | 27 (6) | <0.001<sup>c</sup> |
| Walking time to closest public transport stop (mins) | 6.1 (4.1) | 6.2 (3.0) | 7.2 (7.3) | 7.8 (5.5) | 6.1 (5.6) | 0.22 |
| Number of preferential pedestrian areas | 3 (6) | 3 (1) | 2 (1) | 2 (2) | 3 (2) | 0.001<sup>c</sup> |
| Number of cycling path length (km) | 4.2 (3.1) | 6.7 (0.7) | 1.8 (1.8) | 3.1 (0.6) | 6.7 (0.6) | <0.001<sup>c</sup> |
| Number of interconnected streets >3 | 110 (63) | 133 (50) | 55 (23) | 73 (24) | 133 (57) | <0.001<sup>c</sup> |
| Population density | 8838 (4933) | 11708 (5655) | 5061 (1831) | 7016 (2004) | 12365 (6349) | <0.001<sup>d</sup> |
| Land use mix area coverage | 0.69 (0.11) | 0.72 (0.05) | 0.46 (0.39) | 0.78 (0.29) | 0.71 (0.06) | 0.26 |
| Residential area coverage | 0.33 (0.06) | 0.33 (0.05) | 0.20 (0.17) | 0.36 (0.12) | 0.33 (0.05) | 0.45 |
| Commercial & industrial area coverage | 0.07 (0.08) | 0.18 (0.03) | 0.03 (0.13) | 0.12 (0.10) | 0.18 (0.04) | <0.001<sup>c</sup> |
| Public places area coverage | 0.27 (0.08) | 0.28 (0.02) | 0.21 (0.01) | 0.22 (0.04) | 0.28 (0.02) | <0.001<sup>c</sup> |
| Recreational area coverage | 0.31 (0.11) | 0.32 (0.06) | 0.09 (0.28) | 0.28 (0.23) | 0.33 (0.06) | 0.02<sup>e</sup> |
| Natural area coverage | 0.15 (0.24) | 0 (0.04) | 0.25 (0.09) | 0.30 (0.09) | 0 (0.04) | <0.001<sup>c</sup> |

Except for N, variables are described using the median (IQR).
<sup>a</sup> p-values for High-High vs High-Low vs Low-High vs Low-low
<sup>b</sup> High-High and High-Low were statistically different from Low-Low and Low-High clusters.
<sup>c</sup> High-High and Low-High were statistically different from Low-Low and High-Low clusters.
<sup>d</sup> High-High and Low-High were statistically different from Low-Low and High-Low clusters, but also High-Low statistically different from Low-Low clusters.
<sup>e</sup> Low-High was statistically different from Low-Low clusters.

### Built environment characteristics in MVPA spatial clusters after adjustment for socioeconomic and demographic factors

The characteristics of the built environment among spatial clusters of MVPA adjusted for socioeconomic and demographic factors (Table 2) were similar to the raw MVPA model. Clusters of high MVPA (High-High) presented a higher number of parks (high: 2 *vs* low: 0), public transport stops (high: 22 *vs* low: 12), preferential pedestrian areas (high: 3 *vs* low: 1), cycling pathways (high: 6.7 *vs* low: 2.6 km), interconnected streets (high: 128 *vs* low: 52), population density (high: 8,839 *vs* low: 5,896), and public (high: 0.27 *vs* low: 0.21), and commercial & industrial land use areas (high: 0.20 *vs* low: 0.14) than clusters of low MVPA (Low-Low). High MVPA clusters were also located in areas where the amount of walking time to reach the closest public transport stops was shorter than low MVPA (high 5.6 *vs* low: 8.9 mins). Likewise, clusters of low MVPA showed more natural (low: 0.32 *vs* high: 0) and residential areas (low: 0.37 *vs* high: 0.35) and land use mix area coverage (low: 0.86 *vs* high: 0.73). Socioeconomic and demographic characteristics are described in S3 Table; neighborhood household income was statistically significantly lower in low MVPA clusters.

**Table 2.**
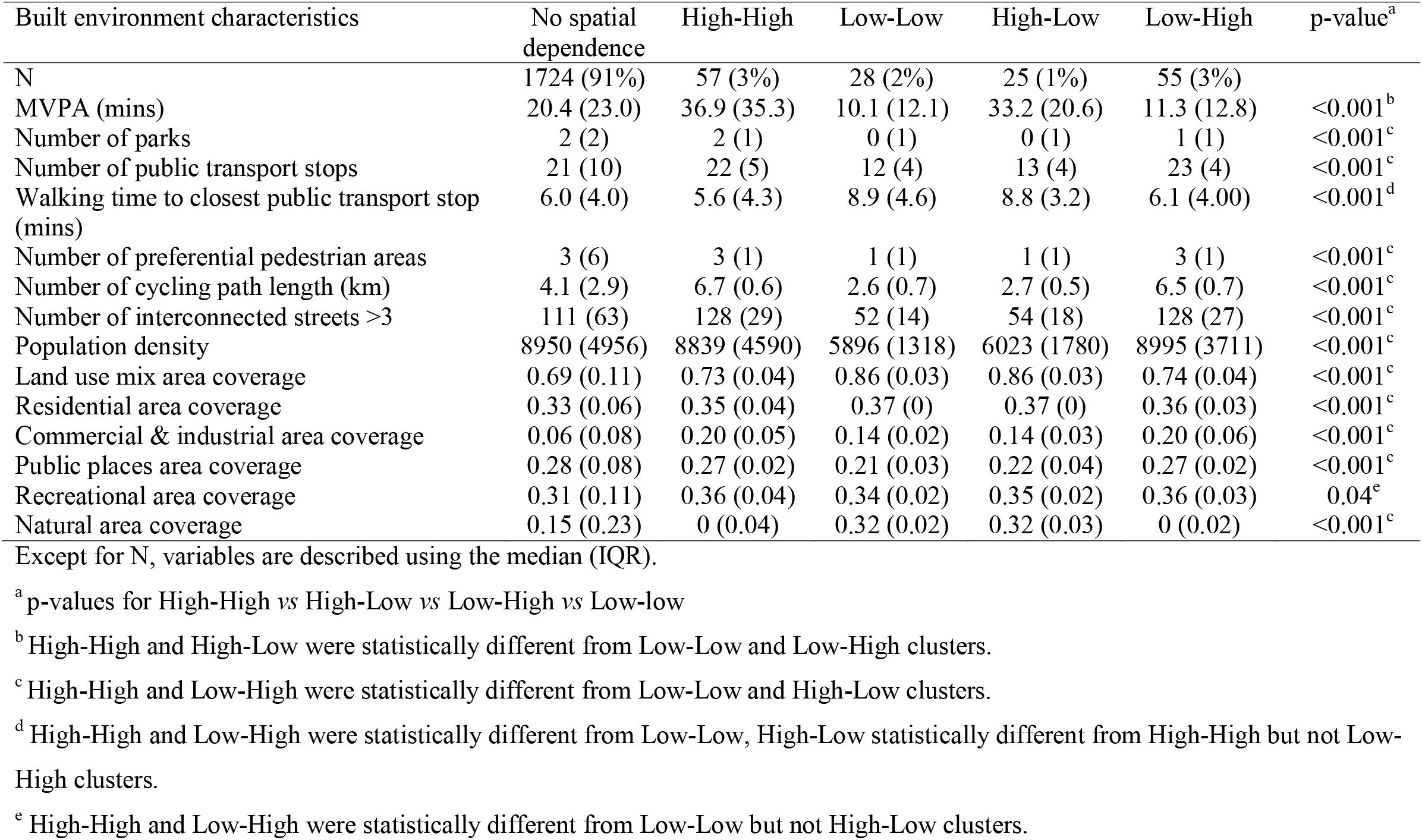
Built environment characteristics in spatial clusters of MVPA adjusted for socioeconomic and demographic factors.

| Built environment characteristics | No spatial dependence | High-High | Low-Low | High-Low | Low-High | p-value<sup>a</sup> |
| --- | --- | --- | --- | --- | --- | --- |
| N | 1724 (91%) | 57 (3%) | 28 (2%) | 25 (1%) | 55 (3%) |  |
| MVPA (mins) | 20.4 (23.0) | 36.9 (35.3) | 10.1 (12.1) | 33.2 (20.6) | 11.3 (12.8) | <0.001<sup>b</sup> |
| Number of parks | 2 (2) | 2 (1) | 0 (1) | 0 (1) | 1 (1) | <0.001<sup>c</sup> |
| Number of public transport stops | 21 (10) | 22 (5) | 12 (4) | 13 (4) | 23 (4) | <0.001<sup>c</sup> |
| Walking time to closest public transport stop (mins) | 6.0 (4.0) | 5.6 (4.3) | 8.9 (4.6) | 8.8 (3.2) | 6.1 (4.00) | <0.001<sup>d</sup> |
| Number of preferential pedestrian areas | 3 (6) | 3 (1) | 1 (1) | 1 (1) | 3 (1) | <0.001<sup>c</sup> |
| Number of cycling path length (km) | 4.1 (2.9) | 6.7 (0.6) | 2.6 (0.7) | 2.7 (0.5) | 6.5 (0.7) | <0.001<sup>c</sup> |
| Number of interconnected streets >3 | 111 (63) | 128 (29) | 52 (14) | 54 (18) | 128 (27) | <0.001<sup>c</sup> |
| Population density | 8950 (4956) | 8839 (4590) | 5896 (1318) | 6023 (1780) | 8995 (3711) | <0.001<sup>c</sup> |
| Land use mix area coverage | 0.69 (0.11) | 0.73 (0.04) | 0.86 (0.03) | 0.86 (0.03) | 0.74 (0.04) | <0.001<sup>c</sup> |
| Residential area coverage | 0.33 (0.06) | 0.35 (0.04) | 0.37 (0) | 0.37 (0) | 0.36 (0.03) | <0.001<sup>c</sup> |
| Commercial & industrial area coverage | 0.06 (0.08) | 0.20 (0.05) | 0.14 (0.02) | 0.14 (0.03) | 0.20 (0.06) | <0.001<sup>c</sup> |
| Public places area coverage | 0.28 (0.08) | 0.27 (0.02) | 0.21 (0.03) | 0.22 (0.04) | 0.27 (0.02) | <0.001<sup>c</sup> |
| Recreational area coverage | 0.31 (0.11) | 0.36 (0.04) | 0.34 (0.02) | 0.35 (0.02) | 0.36 (0.03) | 0.04<sup>e</sup> |
| Natural area coverage | 0.15 (0.23) | 0 (0.04) | 0.32 (0.02) | 0.32 (0.03) | 0 (0.02) | <0.001<sup>c</sup> |

Except for N, variables are described using the median (IQR).
<sup>a</sup> p-values for High-High vs High-Low vs Low-High vs Low-low
<sup>b</sup> High-High and High-Low were statistically different from Low-Low and Low-High clusters.
<sup>c</sup> High-High and Low-High were statistically different from Low-Low and High-Low clusters.
<sup>d</sup> High-High and Low-High were statistically different from Low-Low, High-Low statistically different from High-High but not Low-High clusters.
<sup>e</sup> High-High and Low-High were statistically different from Low-Low but not High-Low clusters.

## Discussion

We observed geographic clusters of high and low MVPA in the urban area of Lausanne. Despite the low number of individuals within clusters, we identified clear differences in the composition of the built environment among clusters, showing that there is a particular underlying composition of the built environment in high MVPA clusters, and a clearly different composition of the built environment in low MVPA clusters. Spatial clusters of high MVPA were located in zones with a higher population density, a better accessibility to parks and to public transportation, more cycling pathways, preferential pedestrian zones, interconnected streets, and public, commercial & industrial zones. As an example, the larger high MVPA cluster (landmark #1 on Fig 1) was clearly located right next to a recreation area equipped with sports facilities on the shores of the lake. The geographic distribution of MVPA was similar for the raw MVPA model and the model adjusted for socioeconomic and demographic factors.

Despite socioeconomic status has been associated as a mediator in the association of the built environment and physical activity [41,42], we did not find clear socioeconomic and demographic differences among clusters of low and high MVPA. However, we did highlight an older population and a higher proportion of non-workers in low raw MVPA clusters, and a lower neighborhood household income in low MVPA clusters after adjustment for socioeconomic and demographic factors. Huang et al. [21], Tamura et al. [19], and Valson et al. [23] also observed inconsistent findings related to socioeconomic determinants and spatial clusters of physical activity. These inconsistencies may be related to our lack of adjustment for social conditions, such as crime rates, pedestrian safety, and harmful land uses [43].

Studies that have explored the association of the built environment with physical activity from a geographic perspective have found mixed evidence [19,20,22]. However, there is an overall agreement that environments that favor walkability, such as higher population density and street connectivity, are associated with higher physical activity levels [18,19,21,23]. In contrast, we did not find a positive association between a higher land use area coverage with the spatial clusters of high physical activity [18,19,23]. This is probably because we were unable to stratify by different types of commercial & industrial land use areas such as food, retails, services, cultural, and physical activity as proposed by Frank et al. [32].

The association between spatial clusters of physical activity and green areas is debatable, with no clear positive association [20,23]. We found that clusters with high MVPA exhibited a higher density of parks and recreational areas but not natural areas. This may highlight important considerations when measuring greenness. As natural areas are usually located at the boundaries of the urban perimeter, the presence of greener areas may not necessarily imply they are adequate or accessible for leisure physical activity or active transportation due to long commuting distances to the destination of individuals [44,45]. Additionally, we hypothesized that city dwellers need a minimum of urban infrastructure or of motivation (i.e. shopping) to move, and that they are not attracted by these natural areas despite their obvious health benefits.

In addition to the above built environmental factors, we also included other determinants, which to our knowledge, are not typically used when analyzing the association between the built environment and spatial clusters of physical activity. We observed that clusters with high MVPA values were located in areas with higher accessibility to cycling pathways and areas that favor pedestrians. Such findings are consistent with other studies not following a spatial methodology [7,44,46]. Similarly, as found in our study, higher accessibility to public transport has also been associated to higher levels of physical activity [7].

Geographic patterns of MVPA on weekdays were similar to the patterns on weekends. However, we observed a higher concentration of individuals in MVPA clusters during weekdays, which may highlight different physical activity interactions of individuals with their built environment depending on the day of the week [47,48]. The comparison of the GGIR algorithm with the PAMPRO methodology to calculate MVPA showed some similarities but also some differences in their MVPA spatial distribution. We decided to use GGIR, but certainly, we could also have chosen PAMPRO for our main analysis. Further studies are needed to validate which method is best suited to measure physical activity using accelerometry data.

Interestingly, we observed similarities between the geographic distribution of MVPA and the distribution of BMI observed in a previous study of the same urban area [36]: one cluster of low MVPA overlaps a cluster of high BMI and a cluster of high MVPA overlaps a cluster of low BMI. This finding is aligned with another study that has found overlapping clusters of BMI and physical activity [49] and could be of great help as a measure of the success of policies promoting healthier built environments.

### Limitations

Our study has several limitations. First, we were not able to assess the working environment nor to track the exact location where people performed physical activity, which would be useful to precisely pinpoint what type of built environment characterized best areas where physical activity was performed [50,51]. Second, since this is an observational study, our findings may suffer from reverse causality, and it is possible that individuals selected their place of residence based on neighborhood characteristics that favor their lifestyles [9]. However, evidence shows that the built environment influences physical activity independently of residential self-selection [10]. Third, the size of the population investigated is moderate (N=1,889), and thus there is a low number of individuals to constitute the spatial clusters observed, resulting in a limited statistical power. Fourth, we did not differentiate between transportation and recreational MVPA, and such a distinction could have been of interest to identify how the built environment affects different practices of physical activity [52]. Fifth, our results may not be entirely generalizable to other locations as built environment characteristics may differ.

### Strengths

To the best of our knowledge, this is the first study that assesses spatial dependence of objectively measured MVPA using individual geolocated data, making our study less prone to bias [32,53]. We used additional variables (i.e. accessibility to cycling pathways, pedestrian areas, and public transport) that are not usually assessed in the study of spatial patterns of physical activity and the built environment, and we considered multiple dimensions of the neighborhood environment [54]. We showed that this inclusion brings a new perspective and evidence for such association. Additionally, we assessed individual spatial clusters of MVPA, which allowed us to evaluate the spatial dependence of MVPA on a local scale.

### Impact for public health policy

Our findings highlight the utility of spatial analysis to explore the influence the characteristics of the built environment have on physical activity and to identify populations at risk, which could be useful for the development of public health policies related to urban planning. Evidence on the success of initiatives modifying the built environment infrastructure to improve physical activity levels is promising [55]. Therefore, policymakers should be encouraged to favor the development of public built environment interventions that promote healthier behaviors -and indirectly more ecological cities-, such as increasing the availability of parks and cycling pathways and facilitating access to public transport and pedestrian areas.

## Conclusions

Although with a low number of individuals, geographic clusters of high MVPA were detected in urban areas where specific compositions of the built environment favors physical activity. Adjustment for socioeconomic and demographic factors did not impact the geographic patterns or built environment characteristics of the clusters. Favoring the planning of urban environments promoting physical activity and active lifestyles should be considered when developing public policies.

## Data Availability

Due to the sensitivity of geolocated data and the lack of consent for online posting, individual data cannot be made accessible. Only metadata can be obtained from the authors upon request. Requests can also be performed via the study website www.colaus-psycolaus.ch.

## Acknowledgments

None.

## Supporting information

**S1 Fig. Built environmental characteristics of the Lausanne urban area**.

**S2 Fig. Spatial clusters of raw MVPA using Local Moran’s I statistics and the PAMPRO algorithm**.

**S3 Fig. Spatial clusters of raw MVPA for weekdays (a) and weekends (b) using Local Moran’s I statistics**.

**S1 Appendix. Regression results of MVPA adjusted for socioeconomic and demographic factors**.

**S4 Fig. Spatial clusters of raw MVPA within spatial lags of 400 (a), 600 (b), 1000 (c), and 1200 m (d) using Local Moran’s I statistics**.

**S1 Table. Comparison of population and built environment characteristics between included and removed individuals**.

**S2 Table. Population socioeconomic and demographic characteristics of raw MVPA spatial clusters**.

**S3 Table. Population socioeconomic and demographic characteristics of adjusted MVPA spatial clusters**.

## References

1. Lee I-M, Shiroma EJ, Lobelo F, Puska P, Blair SN, Katzmarzyk PT. Effect of physical inactivity on major non-communicable diseases worldwide: an analysis of burden of disease and life expectancy. The Lancet. 2012;380: 219–229. doi:10.1016/S0140-6736(12)61031-9

2. WHO. Global action plan on physical activity 2018–2030: more active people for a healthier world. Geneva: World Health Organization; 2018.

3. WHO. Global recommendations on physical activity for health. Geneva: World Health Organization; 2010.

4. Guthold R, Stevens GA, Riley LM, Bull FC. Worldwide trends in insufficient physical activity from 2001 to 2016: a pooled analysis of 358 population-based surveys with 1·9 million participants. The Lancet Global Health. 2018;6: e1077–e1086. doi:10.1016/S2214-109X(18)30357-7

5. Bauman AE, Reis RS, Sallis JF, Wells JC, Loos RJ, Martin BW. Correlates of physical activity: why are some people physically active and others not? The Lancet. 2012;380: 258–271. doi:10.1016/S0140-6736(12)60735-1

6. Trost SG, Owen N, Bauman AE, Sallis JF, Brown W. Correlates of adults’ participation in physical activity: review and update. Medicine & Science in Sports & Exercise. 2002;34: 1996–2001.

7. Sallis James F., Floyd Myron F., Rodríguez Daniel A., Saelens Brian E. Role of Built Environments in Physical Activity, Obesity, and Cardiovascular Disease. Circulation. 2012;125: 729–737. doi:10.1161/CIRCULATIONAHA.110.969022

8. WHO. Towards more physical activity in cities: Transforming public spaces to promote physical activity - a key contributor to achieving the Sustainable Development Goals in Europe. Copenhagen: World Health Organization, Regional Office for Europe; 2017.

9. Kärmeniemi M, Lankila T, Ikäheimo T, Koivumaa-Honkanen H, Korpelainen R. The Built Environment as a Determinant of Physical Activity: A Systematic Review of Longitudinal Studies and Natural Experiments. Ann Behav Med. 2018;52: 239–251. doi:10.1093/abm/kax043

10. McCormack GR, Shiell A. In search of causality: a systematic review of the relationship between the built environment and physical activity among adults. International Journal of Behavioral Nutrition and Physical Activity. 2011;8: 125. doi:10.1186/1479-5868-8-125

11. Smith M, Hosking J, Woodward A, Witten K, MacMillan A, Field A, et al. Systematic literature review of built environment effects on physical activity and active transport – an update and new findings on health equity. International Journal of Behavioral Nutrition and Physical Activity. 2017;14: 158. doi:10.1186/s12966-017-0613-9

12. Mena C, Fuentes E, Ormazábal Y, Triana J, Palomo I. Spatial distribution and physical activity: implications for prevention of cardiovascular diseases. Sport Sci Health. 2017;13: 9–16. doi:10.1007/s11332-017-0349-6

13. Rahnama MR, Shaddel L. Urban Green Space Is Spatially Associated with Cardiovascular Disease Occurrence in Women of Mashhad: a Spatial Analysis of Influential Factors on their Presence in Urban Green Spaces. J Urban Health. 2019;96: 653–668. doi:10.1007/s11524-019-00373-1

14. Auchincloss AH, Gebreab SY, Mair C, Roux AVD. A Review of Spatial Methods in Epidemiology, 2000–2010. Annu Rev Public Health. 2012;33: 107–122. doi:10.1146/annurev-publhealth-031811-124655

15. Greenough PG, Nelson EL. Beyond mapping: a case for geospatial analytics in humanitarian health. Conflict and Health. 2019;13: 50. doi:10.1186/s13031-019-0234-9

16. An R, Li X, Jiang N. Geographical Variations in the Environmental Determinants of Physical Inactivity among U.S. Adults. International Journal of Environmental Research and Public Health. 2017;14: 1326. doi:10.3390/ijerph14111326

17. Lee KH, Dvorak RG, Schuett MA, van Riper CJ. Understanding spatial variation of physical inactivity across the continental United States. Landscape and Urban Planning. 2017;168: 61–71. doi:10.1016/j.landurbplan.2017.09.020

18. Mayne DJ, Morgan GG, Jalaludin BB, Bauman AE. The contribution of area-level walkability to geographic variation in physical activity: a spatial analysis of 95,837 participants from the 45 and Up Study living in Sydney, Australia. Population Health Metrics. 2017;15: 38. doi:10.1186/s12963-017-0149-x

19. Tamura K, Puett RC, Hart JE, Starnes HA, Laden F, Troped PJ. Spatial clustering of physical activity and obesity in relation to built environment factors among older women in three U.S. states. BMC Public Health. 2014;14: 1322. doi:10.1186/1471-2458-14-1322

20. Wang J, Lee K, Kwan M-P. Environmental Influences on Leisure-Time Physical Inactivity in the U.S.: An Exploration of Spatial Non-Stationarity. ISPRS International Journal of Geo-Information. 2018;7: 143. doi:10.3390/ijgi7040143

21. Huang L, Stinchcomb DG, Pickle LW, Dill J, Berrigan D. Identifying Clusters of Active Transportation Using Spatial Scan Statistics. American Journal of Preventive Medicine. 2009;37: 157–166. doi:10.1016/j.amepre.2009.04.021

22. Schuurman N, Peters PA, Oliver LN. Are Obesity and Physical Activity Clustered? A Spatial Analysis Linked to Residential Density. Obesity. 2009;17: 2202–2209. doi:https://doi.org/10.1038/oby.2009.119

23. Valson JS, Kutty VR, Soman B, Jissa VT. Spatial Clusters of Diabetes and Physical Inactivity: Do Neighborhood Characteristics in High and Low Clusters Differ? Asia Pac J Public Health. 2019;31: 612–621. doi:10.1177/1010539519879322

24. Houston D. Implications of the modifiable areal unit problem for assessing built environment correlates of moderate and vigorous physical activity. Applied Geography. 2014;50: 40–47. doi:10.1016/j.apgeog.2014.02.008

25. Kwan M-P. The Uncertain Geographic Context Problem. Annals of the Association of American Geographers. 2012;102: 958–968. doi:10.1080/00045608.2012.687349

26. Firmann M, Mayor V, Vidal PM, Bochud M, Pécoud A, Hayoz D, et al. The CoLaus study: a population-based study to investigate the epidemiology and genetic determinants of cardiovascular risk factors and metabolic syndrome. BMC Cardiovascular Disorders. 2008;8: 6. doi:10.1186/1471-2261-8-6

27. Gubelmann C, Vollenweider P, Marques-Vidal P. Of weekend warriors and couch potatoes: Socio-economic determinants of physical activity in Swiss middle-aged adults. Preventive Medicine. 2017;105: 350–355. doi:10.1016/j.ypmed.2017.10.016

28. Esliger DW, Rowlands AV, Hurst TL, Catt M, Murray P, Eston RG. Validation of the GENEA Accelerometer. Med Sci Sports Exerc. 2011;43: 1085–1093. doi:10.1249/MSS.0b013e31820513be

29. Migueles JH, Rowlands AV, Huber F, Sabia S, Hees VT van. GGIR: A Research Community– Driven Open Source R Package for Generating Physical Activity and Sleep Outcomes From Multi-Day Raw Accelerometer Data. JMPB. 2019;2: 188–196. doi:10.1123/jmpb.2018-0063

30. Tom White. Thomite/pampro v0.4.0. Zenodo; 2018. doi:10.5281/zenodo.1187043

31. Dillon CB, Fitzgerald AP, Kearney PM, Perry IJ, Rennie KL, Kozarski R, et al. Number of Days Required to Estimate Habitual Activity Using Wrist-Worn GENEActiv Accelerometer: A Cross-Sectional Study. PLOS ONE. 2016;11: e0109913. doi:10.1371/journal.pone.0109913

32. Frank LD, Schmid TL, Sallis JF, Chapman J, Saelens BE. Linking objectively measured physical activity with objectively measured urban form: Findings from SMARTRAQ. American Journal of Preventive Medicine. 2005;28: 117–125. doi:10.1016/j.amepre.2004.11.001

33. Tobler W. Three Presentations on Geographical Analysis and Modeling: Non-Isotropic Geographic Modeling; Speculations on the Geometry of Geography; and Global Spatial Analysis. UC Santa Barbara: National Center for Geographic Information and Analysis. 1993.

34. Anselin L. Local Indicators of Spatial Association—LISA. Geographical Analysis. 1995;27: 93– 115. doi:10.1111/j.1538-4632.1995.tb00338.x

35. Moran PAP. Notes on Continuous Stochastic Phenomena. Biometrika. 1950;37: 17–23. doi:10.2307/2332142

36. Joost S, Duruz S, Marques-Vidal P, Bochud M, Stringhini S, Paccaud F, et al. Persistent spatial clusters of high body mass index in a Swiss urban population as revealed by the 5-year GeoCoLaus longitudinal study. BMJ Open. 2016;6: e010145. doi:10.1136/bmjopen-2015-010145

37. R Core Team. R: A language and environment for statistical computing. R Foundation for Statistical Computing, Vienna, Austria; 2020.

38. Xun Li. rgeoda: R library for spatial data analysis. R package version 0.0.4. 2019.

39. Edzer Pebesma. Simple Features for R: Standardized Support for Spatial Vector Data. R package version 0.9-1. 2018.

40. Hadley Wickham. ggplot2: Elegant Graphics for Data Analysis. R package version 3.3.2. 2016.

41. Koohsari MJ, Hanibuchi T, Nakaya T, Shibata A, Ishii K, Liao Y, et al. Associations of Neighborhood Environmental Attributes with Walking in Japan: Moderating Effects of Area-Level Socioeconomic Status. J Urban Health. 2017;94: 847–854. doi:10.1007/s11524-017-0199-1

42. Sugiyama T, Howard NJ, Paquet C, Coffee NT, Taylor AW, Daniel M. Do Relationships Between Environmental Attributes and Recreational Walking Vary According to Area-Level Socioeconomic Status? J Urban Health. 2015;92: 253–264. doi:10.1007/s11524-014-9932-1

43. Weiss CC, Purciel M, Bader M, Quinn JW, Lovasi G, Neckerman KM, et al. Reconsidering Access: Park Facilities and Neighborhood Disamenities in New York City. J Urban Health. 2011;88: 297–310. doi:10.1007/s11524-011-9551-z

44. Mäki-Opas Te, Borodulin K, Valkeinen H, Stenholm S, Kunst AE, Abel T, et al. The contribution of travel-related urban zones, cycling and pedestrian networks and green space to commuting physical activity among adults – a cross-sectional population-based study using geographical information systems. BMC Public Health. 2016;16: 760. doi:10.1186/s12889-016-3264-x

45. Garden FL, Jalaludin BB. Impact of Urban Sprawl on Overweight, Obesity, and Physical Activity in Sydney, Australia. J Urban Health. 2009;86: 19–30. doi:10.1007/s11524-008-9332-5

46. Charreire H, Weber C, Chaix B, Salze P, Casey R, Banos A, et al. Identifying built environmental patterns using cluster analysis and GIS: Relationships with walking, cycling and body mass index in French adults. International Journal of Behavioral Nutrition and Physical Activity. 2012;9: 59. doi:10.1186/1479-5868-9-59

47. Cerin E, Mitáš J, Cain KL, Conway TL, Adams MA, Schofield G, et al. Do associations between objectively-assessed physical activity and neighbourhood environment attributes vary by time of the day and day of the week? IPEN adult study. International Journal of Behavioral Nutrition and Physical Activity. 2017;14: 34. doi:10.1186/s12966-017-0493-z

48. Clary C, Lewis D, Limb ES, Nightingale CM, Ram B, Rudnicka AR, et al. Weekend and weekday associations between the residential built environment and physical activity: Findings from the ENABLE London study. PLOS ONE. 2020;15: e0237323. doi:10.1371/journal.pone.0237323

49. Michimi A, Wimberly MC. Spatial Patterns of Obesity and Associated Risk Factors in the Conterminous U.S. American Journal of Preventive Medicine. 2010;39: e1–e12. doi:10.1016/j.amepre.2010.04.008

50. Carlson JA, Frank LD, Ulmer J, Conway TL, Saelens BE, Cain KL, et al. Work and Home Neighborhood Design and Physical Activity. Am J Health Promot. 2018;32: 1723–1729. doi:10.1177/0890117118768767

51. Holliday KM, Howard AG, Emch M, Rodríguez DA, Rosamond WD, Evenson KR. Where Are Adults Active? An Examination of Physical Activity Locations Using GPS in Five US Cities. J Urban Health. 2017;94: 459–469. doi:10.1007/s11524-017-0164-z

52. Saelens BE, Handy SL. Built Environment Correlates of Walking: A Review. Medicine & Science in Sports & Exercise. 2008;40: S550. doi:10.1249/MSS.0b013e31817c67a4

53. Limb ES, Ahmad S, Cook DG, Kerry SM, Ekelund U, Whincup PH, et al. Measuring change in trials of physical activity interventions: a comparison of self-report questionnaire and accelerometry within the PACE-UP trial. International Journal of Behavioral Nutrition and Physical Activity. 2019;16: 10. doi:10.1186/s12966-018-0762-5

54. Schüle SA, Bolte G. Interactive and Independent Associations between the Socioeconomic and Objective Built Environment on the Neighbourhood Level and Individual Health: A Systematic Review of Multilevel Studies. PLOS ONE. 2015;10: e0123456. doi:10.1371/journal.pone.0123456

55. Mayne SL, Auchincloss AH, Michael YL. Impact of Policy and Built Environment Changes on Obesity-related Outcomes: A Systematic Review of Naturally-Occurring Experiments. Obes Rev. 2015;16: 362–375. doi:10.1111/obr.12269

