## Supplementary material for "Geographic clusters of objectively measured physical activity and the characteristics of their built environment in a Swiss urban area": S1 Fig

**SUPPLEMENTARY INFORMATION**

**Urban area of Lausanne**

**S1 Fig. Built environmental characteristics of the Lausanne urban area.**

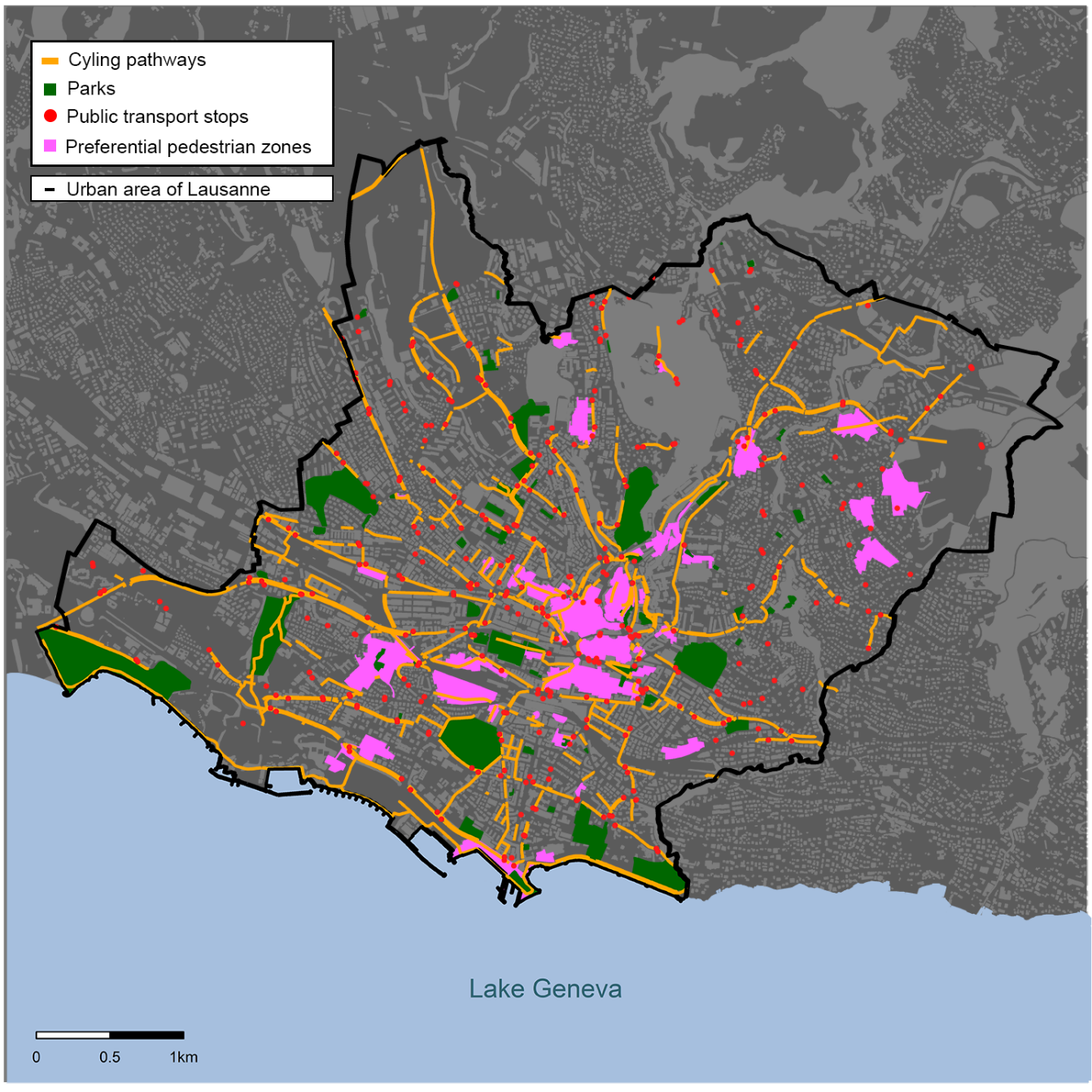
