## Supplementary material for "Geographic clusters of objectively measured physical activity and the characteristics of their built environment in a Swiss urban area": S2 Fig

**Spatial distribution of raw MVPA (PAMPRO algorithm)**

**S2 Fig. Spatial clusters of raw MVPA using Local Moran’s I statistics and the PAMPRO algorithm.** Statistical significance is assessed based on an α threshold of p<0.05 within a spatial lag of 800 m. Dark-green dots indicate individuals with high MVPA values surrounded by neighbors also showing high MVPA values; red dots indicate individuals with low MVPA surrounded by neighbors with low MVPA values; light-green dots indicate individuals with high MVPA values surrounded by neighbors showing low MVPA values; pink dots indicate individuals with low MVPA values surrounded by neighbors with high MVPA values; white dots indicate individuals whose MVPA values are randomly distributed in the geographic space. Landmarks (1-4) are shown to facilitate the description and interpretation of the results.

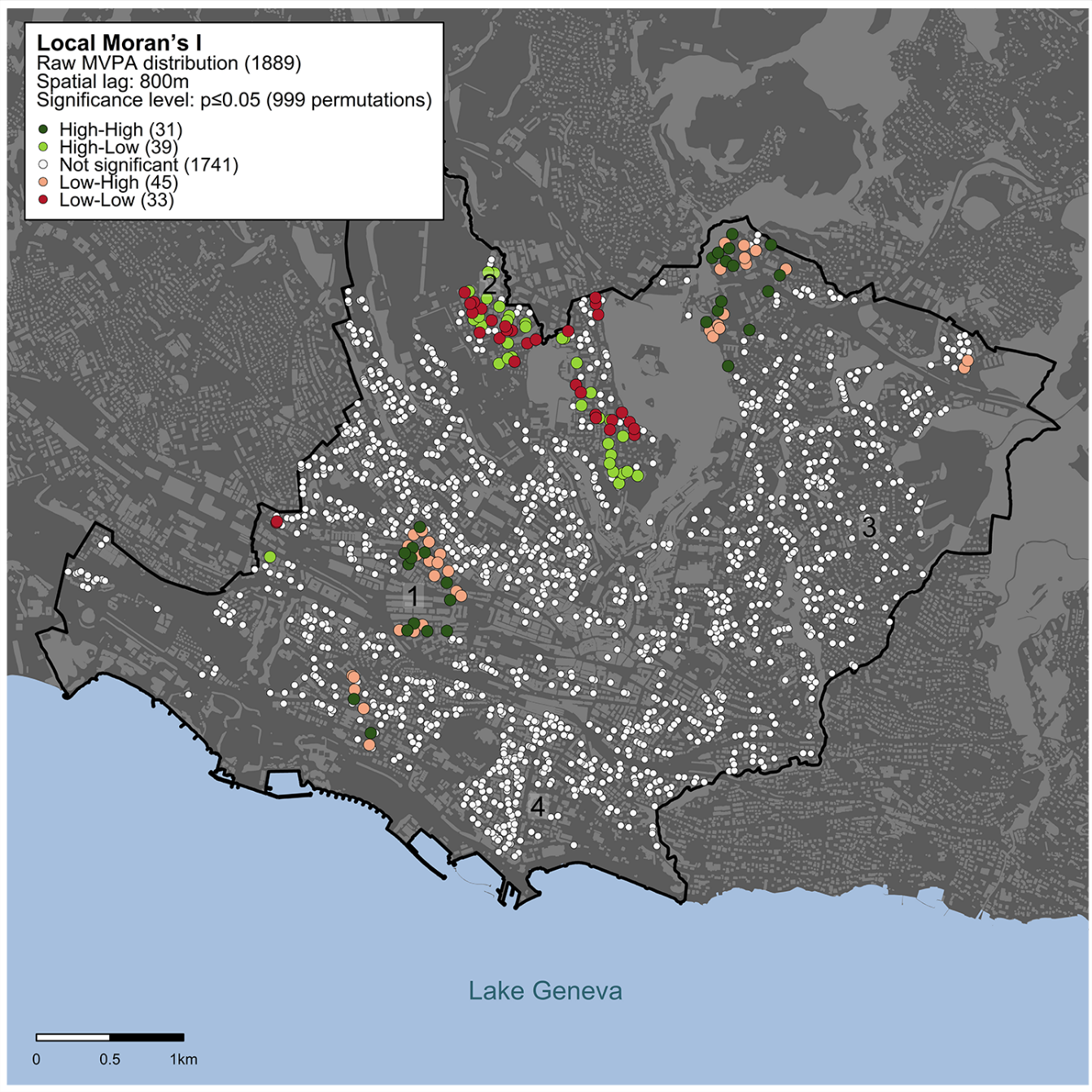
