## Supplementary material for "Geographic clusters of objectively measured physical activity and the characteristics of their built environment in a Swiss urban area": S1 Appendix

**S1 Appendix. Regression results of MVPA adjusted for socioeconomic and demographic factors.**

| Variable | Coefficient | Confidence Intervals (95%) | Standard Error | P-value |
| --- | --- | --- | --- | --- |
| Intercept | 11.84 | 10.83, 12.85 | 0.56 | <0.001 |
| Age (years) | -0.07 | -0.09, -0.06 | 0.01 | <0.001 |
| Neighborhood household income (USD) | 0.003 | -0.01, 0.01 | 0.01 | 0.55 |
| BMI (kg/m^2^) | -0.07 | -0.09, -0.05 | 0.01 | <0.001 |
| Men vs women | -0.08 | -0.27, 0.09 | 0.10 | 0.38 |
| Medium vs low education level | -0.25 | -0.46, 0.04 | 0.11 | 0.05 |
| High vs low education level | -0.28 | -0.53, 0.02 | 0.13 | 0.05 |
| Married vs single | 0.05 | -0.19, 0.30 | 0.14 | 0.73 |
| Divorced vs single | -0.06 | -0.35, 0.22 | 0.16 | 0.65 |
| Widowed vs single | -0.42 | -0.84, -0.01 | 0.21 | 0.04 |
| White vs non-white | -0.08 | -0.41, 0.24 | 0.17 | 0.61 |
| Medium vs low job status | -0.17 | -0.45, 0.11 | 0.16 | 0.28 |
| High vs low job status | -0.41 | -0.77, -0.04 | 0.20 | 0.04 |
| Not working vs low job status | -0.32 | -0.58, -0.05 | 0.15 | 0.03 |
