## Supplementary material for "Geographic clusters of objectively measured physical activity and the characteristics of their built environment in a Swiss urban area": S4 Fig

**Spatial distribution of MVPA using different spatial lags (400, 600, 1000, and 1200 m)**

**S4 Fig. Spatial clusters of raw MVPA within spatial lags of 400 (a), 600 (b), 1000 (c), and 1200 m (d) using Local Moran’s I statistics.** Statistical significance is assessed based on an α threshold of p<0.05. Dark-green dots indicate individuals with high MVPA values surrounded by neighbors also showing high MVPA values; red dots indicate individuals with low MVPA surrounded by neighbors with low MVPA values; light-green dots indicate individuals with high MVPA values surrounded by neighbors showing low MVPA values; pink dots indicate individuals with low MVPA values surrounded by neighbors with high MVPA values; white dots indicate individuals whose MVPA values are randomly distributed in the geographic space. Landmarks (1-4) are shown to facilitate the description and interpretation of the results.

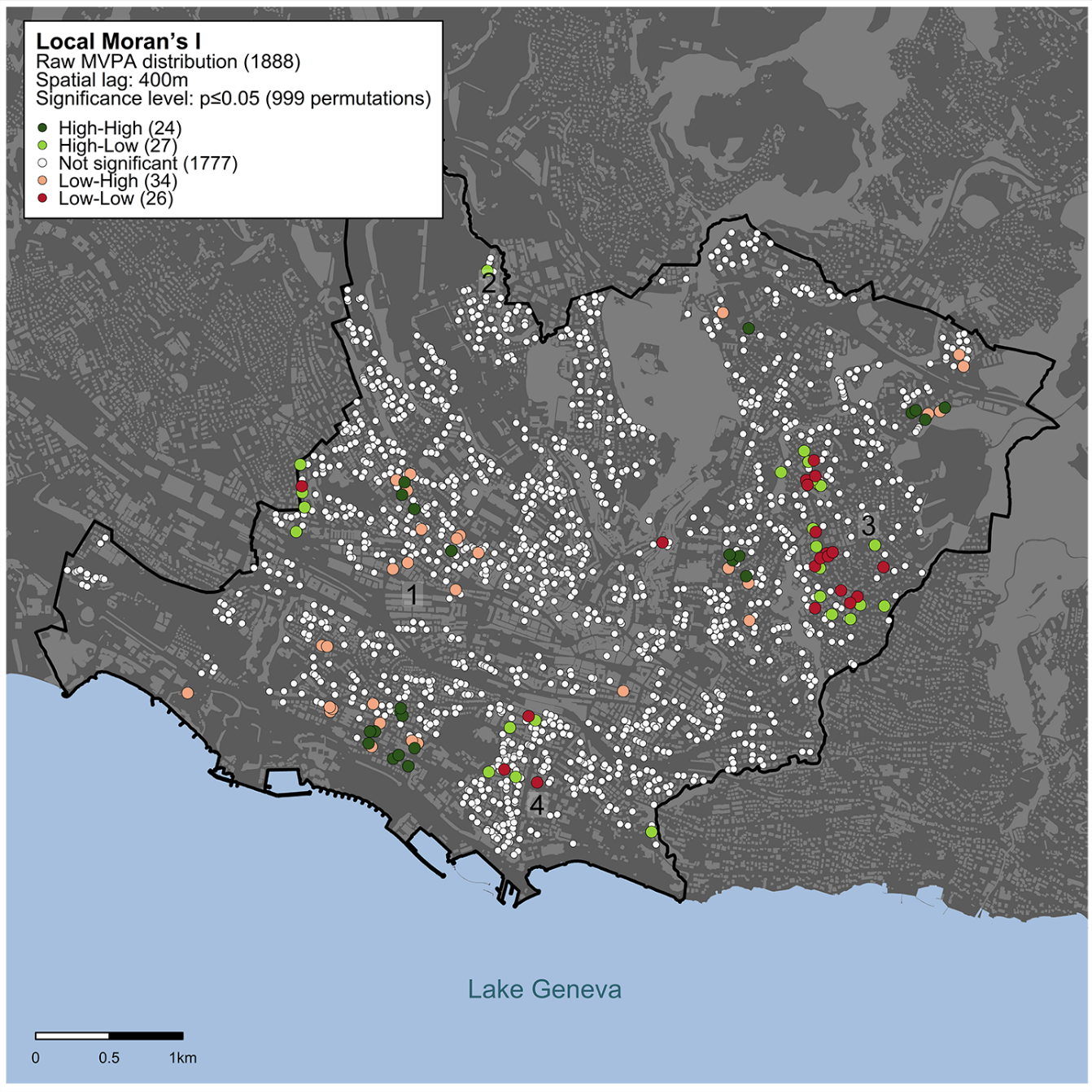

a

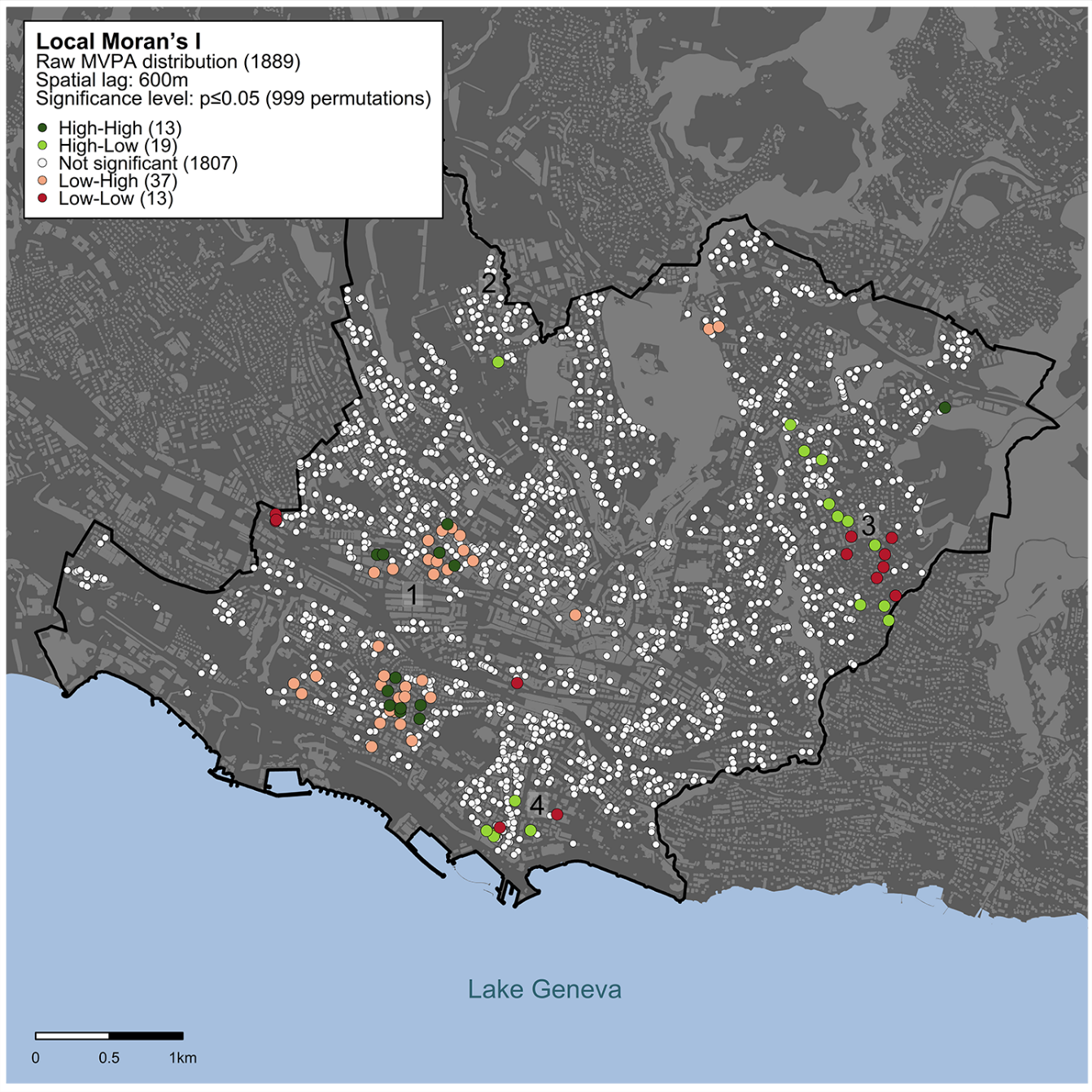

b

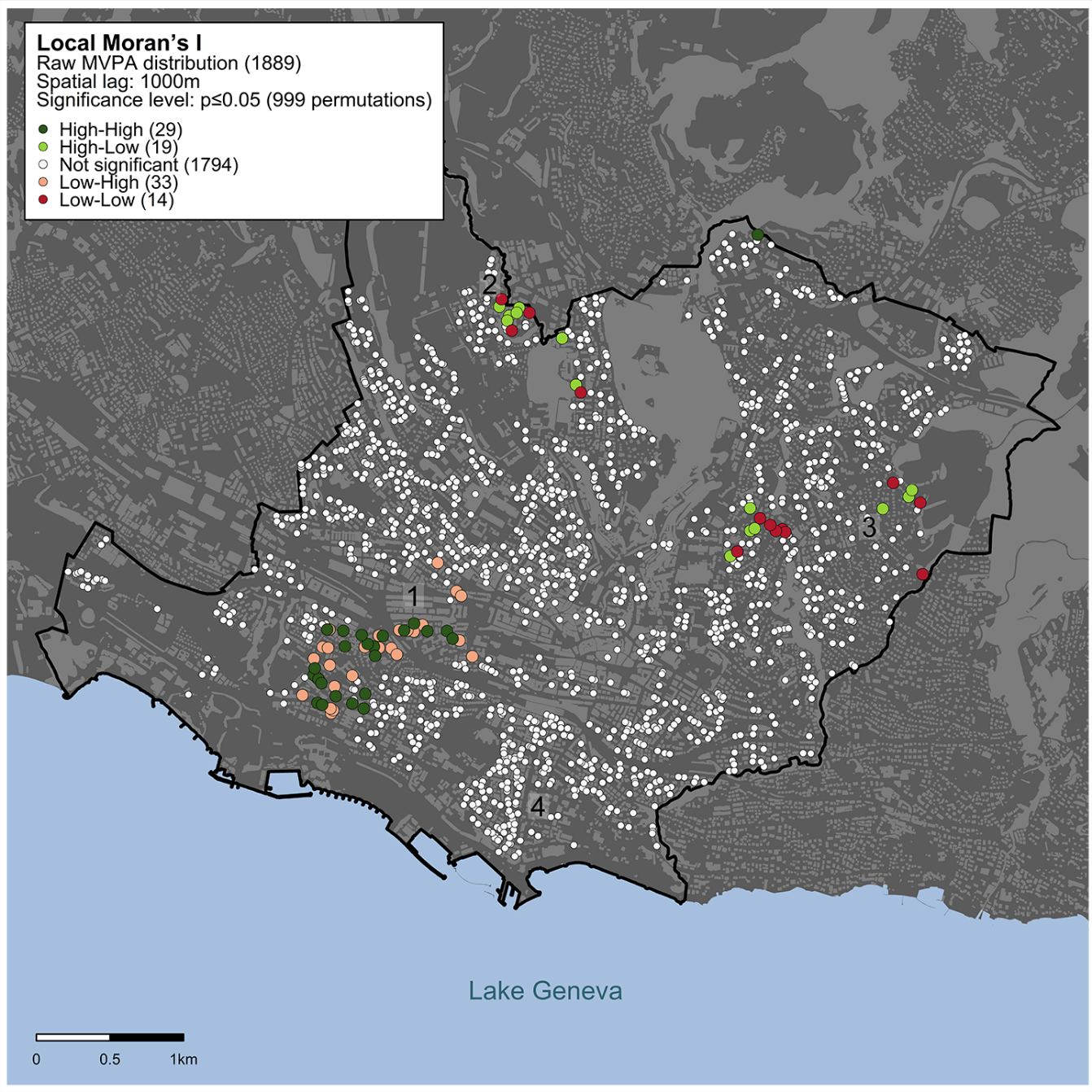

c

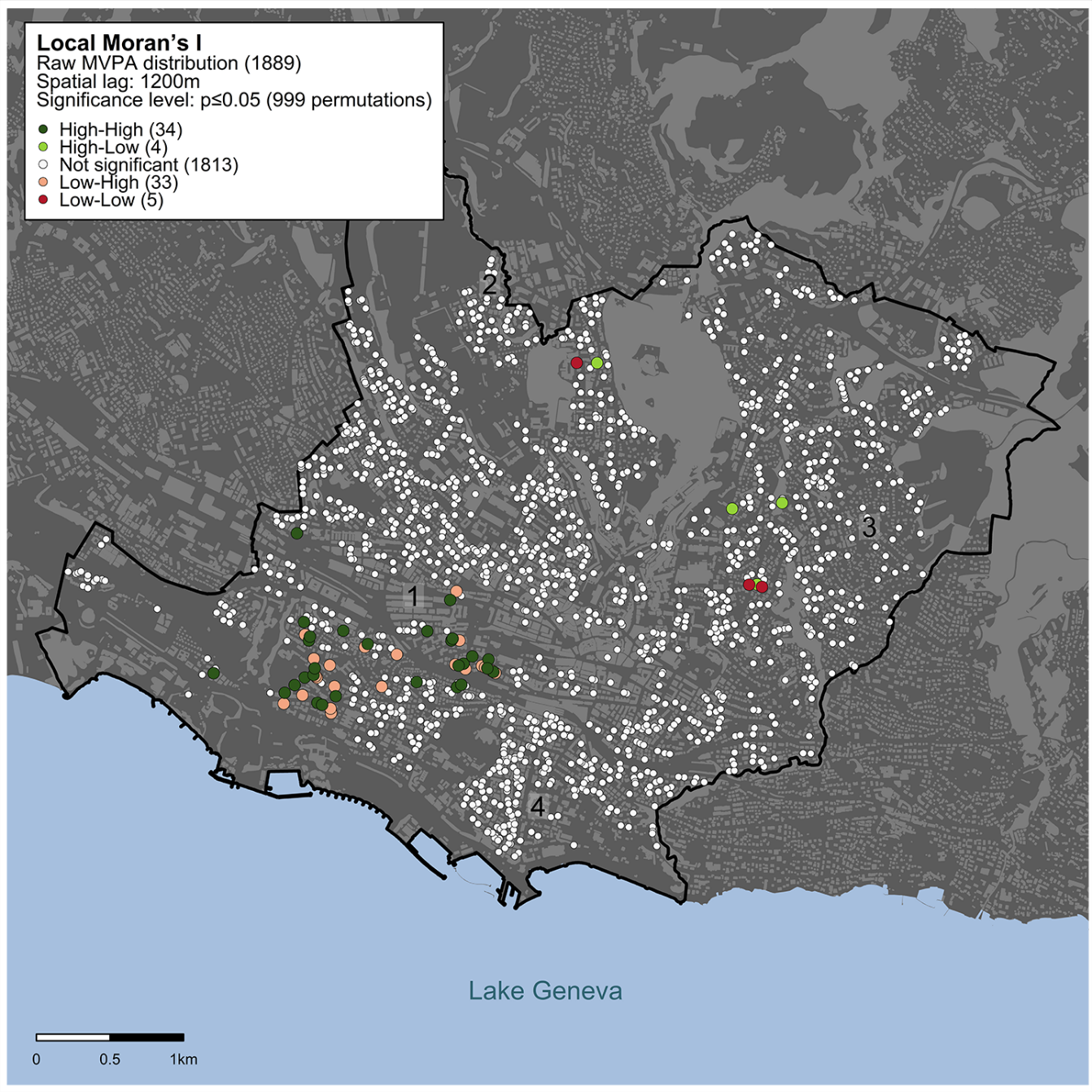

d
