## Supplementary material for "Geographic clusters of objectively measured physical activity and the characteristics of their built environment in a Swiss urban area": S1 Table

**S1 Table. Comparison of population and built environment characteristics between included and removed individuals.**

|  | Population included in the analysis | Population excluded from the analysis^a^ |  |
| --- | --- | --- | --- |
|  | Median (IQR) or  Frequencies (%) | Median (IQR) or  Frequencies (%) | p-values |
| Population Characteristics |  |  |  |
| N | 1889 | 352 | - |
| MVPA (mins per day) | 20.3 (23.0) | 19.9 (26.1) | 0.96 |
| Age (years) | 63 (16) | 60 (16) | 0.01 |
| Household Income (USD) | 56253 (13093) | 55970 (13152) | 0.77 |
| BMI (kg/m^2^) | 25.8 (5.8) | 26.4 (6.7) | 0.07 |
| Women | 1409 (55%) | 215 (61%) | 0.07 |
| Civil status |  |  | 0.03 |
| Single | 282 (15%) | 46 (15%) |  |
| Married | 1069 (56%) | 162 (53%) |  |
| Divorced | 412 (22%) | 66 (21%) |  |
| Widowed | 126 (7%) | 35 (11%) |  |
| White | 1742 (92%) | 318 (90%) | 0.28 |
| Education |  |  | 0.02 |
| Low | 986 (52%) | 205 (59%) |  |
| Medium | 495 (26%) | 67 (19%) |  |
| High | 408 (22%) | 77 (22%) |  |
| Job status |  |  | 0.24 |
| Low | 438 (23%) | 54 (25%) |  |
| Medium | 322 (17%) | 30 (14%) |  |
| High | 188 (10%) | 29 (14%) |  |
| Not working | 941 (50%) | 100 (47%) |  |
| Built environmental characteristics |  |  |  |
| Number of parks | 2 (2) | 2 (2) | 0.94 |
| Number of public transport stops | 20 (10) | 20 (22) | 0.57 |
| Walking time to closest public transport stop (mins) | 6.1 (4.1) | 6.0 (4.3) | 0.48 |
| Number of preferential pedestrian areas | 3 (6) | 3 (5) | 0.58 |
| Number of cycling path length (km) | 4.2 (3.2) | 4.5 (3.1) | 0.46 |
| Number of interconnected streets >3 | 111 (63) | 112 (65) | 0.60 |
| Population density | 8824 (4960) | 8956 (5039) | 0.49 |
| Land use mix area coverage | 0.70 (0.11) | 0.69 (0.11) | 0.98 |
| Residential area coverage | 0.33 (0.06) | 0.33 (0.07) | 0.57 |
| Commercial **&** industrial area coverage | 0.07 (0.09) | 0.07 (0.09) | 0.95 |
| Public places area coverage | 0.27 (0.08) | 0.28 (0.07) | 0.65 |
| Recreational area coverage | 0.31 (0.10) | 0.31 (0.10) | 0.92 |
| Natural area coverage | 0.15 (0.24) | 0.15 (0.24) | 0.81 |

^a^Population with missing covariates or invalid accelerometry data only. Population outside the urban area of Lausanne or without geolocation data (n=726) are not described in the table as built environment characteristics may not be comparable.
