## Supplementary material for "Geographic clusters of objectively measured physical activity and the characteristics of their built environment in a Swiss urban area": S2 Table

**S2 Table**. **Population socioeconomic and demographic characteristics of raw MVPA spatial clusters.**

| Socioeconomic characteristics | No spatial dependence | High-High | Low-Low | High-Low | Low-High | p-value^a^ |
| --- | --- | --- | --- | --- | --- | --- |
|  | Median (IQR) or  Frequencies (%) | Median (IQR) or  Frequencies (%) | Median (IQR) or  Frequencies (%) | Median (IQR) or  Frequencies (%) | Median (IQR) or  Frequencies (%) |  |
| N | 1793 (95%) | 27 (1%) | 13 (1%) | 14 (1%) | 42 (2%) |  |
| Age (years) | 63 (16) | 55 (12) | 73 (16) | 55 (10) | 68 (13) | <0.001 |
| Neighborhood household income (USD) | 56253 (12025) | 56520 (7616) | 65562 (56973) | 55094 (17203) | 56613 (8180) | 0.40 |
| BMI (kg/m^2^) | 25.9 (5.9) | 24.9 (4.4) | 25.4 (6.4) | 24.5 (4.0) | 26.7 (5.5) | 0.31 |
| Women | 986 (55%) | 16 (59%) | 7 (54%) | 12 (86%) | 28 (67%) | 0.28 |
| Civil status |  |  |  |  |  | 0.44 |
| Single | 268 (15%) | 4 (15%) | 4 (31%) | 3 (21%) | 3 (7%) |  |
| Married | 1009 (56%) | 17 (63%) | 8 (61%) | 10 (71%) | 25 (60%) |  |
| Divorced | 395 (22%) | 5 (18%) | 1 (8%) | 1 (8%) | 10 (24%) |  |
| Widowed | 121 (7%) | 1 (4%) | (%) | 0 (0%) | 4 (9%) |  |
| White | 1653 (92%) | 25 (93%) | 13 (100%) | 11 (79%) | 40 (95%) | 0.18 |
| Education |  |  |  |  |  | 0.46 |
| Low | 933 (52%) | 15 (56%) | 5 (38%) | 10 (71%) | 23 (55%) |  |
| Medium | 472 (26%) | 5 (18%) | 5 (38%) | 1 (8%) | 12 (28%) |  |
| High | 388 (22%) | 7 (26%) | 3 (24%) | 3 (21%) | 7 (17%) |  |
| Job status |  |  |  |  |  | <0.001 |
| Low | 417 (23%) | 9 (33%) | 0 (0%) | 7 (50%) | 5 (12%) |  |
| Medium | 309 (17%) | 4 (15%) | 2 (15%) | 3 (21%) | 4 (9%) |  |
| High | 178 (10%) | 6 (22%) | 1 (8%) | 1 (8%) | 2 (5%) |  |
| Not working | 889 (50%) | 8 (30%) | 10 (77%) | 3 (21%) | 31 (74%) |  |

^a^p-values for High-High *vs* High-Low *vs* Low-High *vs* Low-low
