## Supplementary material for "Geographic clusters of objectively measured physical activity and the characteristics of their built environment in a Swiss urban area": S3 Table

**S3** **Table. Population socioeconomic and demographic characteristics of adjusted MVPA spatial clusters.**

| Socioeconomic characteristics | No spatial dependence | High-High | Low-Low | High-Low | Low-High | p-value^a^ |
| --- | --- | --- | --- | --- | --- | --- |
|  | Median (IQR) or  Frequencies (%) | Median (IQR) or  Frequencies (%) | Median (IQR) or  Frequencies (%) | Median (IQR) or  Frequencies (%) | Median (IQR) or  Frequencies (%) |  |
| N | 1724 (91%) | 57 (3%) | 28 (2%) | 25 (1%) | 55 (3%) |  |
| Age (years) | 63 (16) | 62 (21) | 67 (19) | 67 (19) | 62 (18) | 0.97 |
| Neighborhood household income (USD) | 56253 (12309) | 56705 (11627) | 45774 (0) | 45774 (0) | 56705 (12368) | <0.001 |
| BMI (kg/m^2^) | 25.9 (5.9) | 25.8 (6.8) | 25.4 (3.5) | 25.8 (5.0) | 26.0 (5.4) | 0.84 |
| Women | 953 (55%) | 32 (56%) | 17 (61%) | 12 (48%) | 35 (64%) | 0.60 |
| Civil status |  |  |  |  |  | 0.37 |
| Single | 263 (15%) | 7 (12%) | 4 (14%) | 3 (12%) | 5 (9%) |  |
| Married | 968 (56%) | 30 (53%) | 14 (50%) | 17 (68%) | 40 (73%) |  |
| Divorced | 382 (22%) | 13 (23%) | 8 (29%) | 4 (16%) | 5 (9%) |  |
| Widowed | 111 (7%) | 7 (12%) | 1 (7%) | 1 (4%) | 5 (9%) |  |
| White | 1590 (92%) | 54 (95%) | 24 (86%) | 22 (88%) | 52 (95%) | 0.34 |
| Education |  |  |  |  |  | 0.90 |
| Low | 892 (52%) | 33 (58%) | 17 (61%) | 15 (60%) | 29 (52%) |  |
| Medium | 456 (26%) | 12 (21%) | 7 (25%) | 7 (28%) | 13 (24%) |  |
| High | 376 (22%) | 12 (21%) | 4 (14%) | 3 (12%) | 13 (24%) |  |
| Job status |  |  |  |  |  | 0.86 |
| Low | 399 (23%) | 13 (23%) | 6 (21%) | 7 (28%) | 13 (24%) |  |
| Medium | 295 (17%) | 9 (16%) | 5 (18%) | 5 (20%) | 8 (14%) |  |
| High | 174 (10%) | 6 (10%) | 0 (0%) | 2 (8%) | 6 (11%) |  |
| Not working | 856 (50%) | 29 (51%) | 17 (61%) | 11 (44%) | 28 (51%) |  |

^a^p-values for High-High *vs* High-Low *vs* Low-High *vs* Low-low
